# Whole Care Home Testing for Covid-19 in a Local Authority Area in the United Kingdom

**DOI:** 10.1101/2020.08.06.20162859

**Authors:** Anna Starling, Eleanor White, Danny Showell, David Wyllie, Smita Kapadia, Ravi Balakrishnan

## Abstract

**Objectives:** To describe the point prevalence of SARS-CoV-2 in care homes reporting low numbers of cases of COVID-19.

**Design:** A cross-sectional study of care homes, ascertaining perceived disease burden using interviews with care home managers and SARS-CoV-2 RNA detection in residents and staff using nose and throat swabbing.

**Setting:** 15 Care homes in Essex, United Kingdom, all of which had reported either zero or one case of COVID-19 to the Health Protection Team.

**Participants:** 912 residents and staff of care homes were tested. Residents were eligible to be tested regardless of symptoms.

**Main outcome measure:** Detection of SARS-CoV-2 in residents and staff.

**Results:** In the 15 care homes studied, SARS-CoV-2 was detected in 23 (5.2%) of 441 residents. Of these 23, 21/23 (91%) were asymptomatic as reported by the care home managers. SARS-CoV-2 was detected in 8/471 (1.7%) of staff. This differs from that in residents (p=0.003).

**Conclusions:** The study’s findings suggest that symptoms, as reported by care home managers, are an insensitive method of defining the extent of SARS-CoV-2 infection in nursing homes. Viral detection from residents is more common than from staff. Microbiological screening is a more sensitive method for defining the extent of SARS-CoV-2 in care homes than managerial reporting of resident symptoms.

## INTRODUCTION

On the 31^st^ December 2019, a new cluster of pneumonia of unknown aetiology was first reported in Wuhan, China^1^. The cause was later identified as a novel coronavirus. The World Health Organisation named the virus severe acute respiratory syndrome coronavirus 2 (SARS-CoV-2)^2^. The first indigenous case in the United Kingdom (UK) with no direct or indirect travel links was confirmed on 28^th^ February 2020^3^ and the first COVID-19 death on 5^th^ March 2020^4^.

As the epidemic has progressed, there is evidence that residential care environments, including care homes, may be playing an increasing role in transmission and the generation of new cases^5^. As of 18 April, there were: 3,500 care home outbreaks cumulative, reported to Public Health England (PHE) in England, accounting for 22% of the care homes. A point-prevalence study in London of four care homes identified COVID-19 in 40% of residents, 43% of whom were asymptomatic and 18% had atypical symptoms^6^. Recent analysis of whole care home testing data for over 65s in England between May and June 2020 (9081 homes) identified COVID-19 in 3.9% of residents, 80.9% of whom were reported as asymptomatic^7^.

In April and May 2020 Essex County Council and PHE undertook a cross-sectional study in care homes (all ages) reporting low or no COVID-19 related illness among their residents and staff. The primary objective was to determine the proportion of care home residents and care home workers at work in whom SARS-CoV-2 can be detected to increase understanding of the prevalence of COVID-19 in care home settings.

## METHODS

### Home survey

On the week commencing the 26^th^ April 2020, a COVID-19 outbreak had been reported in 23% of care homes in Essex. We identified 23 homes which had reported to the PHE Health Protection team either zero or one COVID-19 case(s). This was an opportunistic and pragmatic approach to intervene early to prevent transmission in homes and was therefore rooted in public health action. 21 of the 23 homes invited agreed to take part. Due to changes in testing capacity 16 homes were included in the final selection. The decision regarding which homes to include was made based on the order they were recruited, with the latter six homes being excluded.

Residents were eligible to be tested for SARs-CoV-2 regardless of symptoms. All care home staff who considered themselves to be fit to work were also eligible to be tested, including bank and agency staff. A PCR-based swab test was used to detect SARS-CoV-2 infection in the upper respiratory tract samples. Swabbing took place at the care homes between the 1^st^ and 14^th^ May 2020, undertaken by a locally commissioned service that comprised trained nurses and health care assistants.

### Microbiological methods

In 14 homes the testing was via the regional Public Health laboratory. Two homes had testing done via a network of dedicated COVID-19 testing laboratories (Pillar 2). The assays used by these laboratories have similar performance and highly specific^8^.

### Questionnaire

Care home managers were interviewed using a structured questionnaire in advance of swabbing to provide information on the total number of staff, residents, confirmed cases and deaths prior to participation. Following swabbing, care home managers were asked to provide basic demographic information for those swabbed (age, sex and ethnicity) and details of any symptomatic residents on the day of swabbing, based on the then UK Government case definition for COVID-19 of a new continuous cough or fever^9^.

### Follow up

All residents and staff in whom SARS-CoV-2 was detected were followed up to understand if they developed symptoms up to 14 days following swabbing. This information was collected from the care home managers by the research team. Staff who were positive but asymptomatic were offered an additional PCR-based swab test ten days after testing to determine if SARS-CoV-2 could still be detected. (Figure 1)

**Figure 1:**
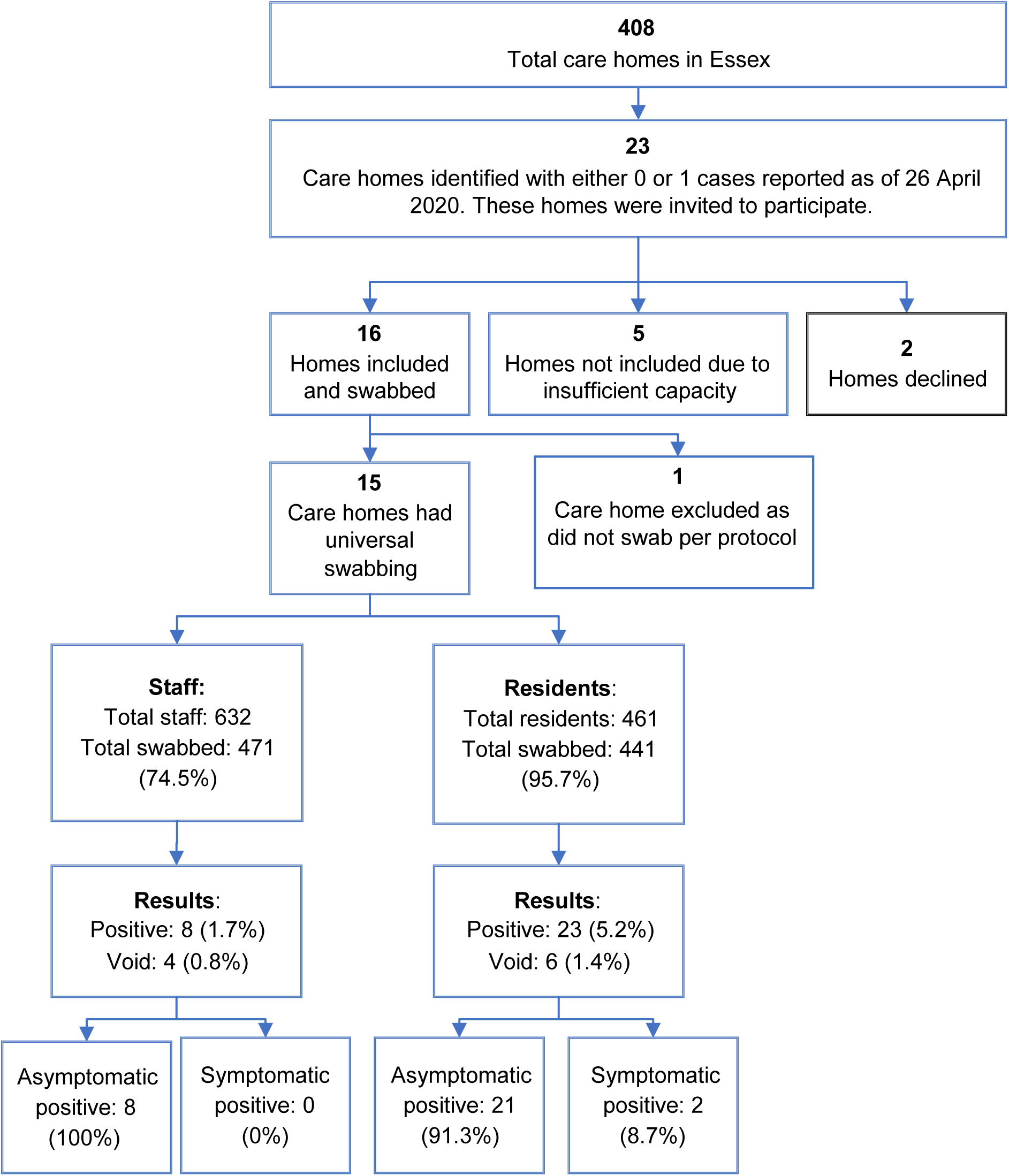
Care Home Testing Process.

### Statistical methods

Results for residents and staff were examined separately. In addition, homes were stratified according to those with no confirmed (with at least one positive test result in staff or residents) or suspected cases, as reported to the research team by the care home manager, prior to the study swabbing. We compared counts of individuals with positive SARS-CoV-2 tests between residents and staff using a x^2^ test with Yates’ correction for Continuity (R 3.3.1).

## RESULTS

The results from 15 care homes were included in the study, the results from the 16th care home were excluded due to selective sampling, as opposed to universal swabbing, of residents. (Figure 1).

The care homes included in the study ranged in size from ten to 67 residents. Three of the homes had no suspected or confirmed clinical cases in residents or staff prior to swab-testing taking place as reported to the research team (Table 1).

**Table 1.**
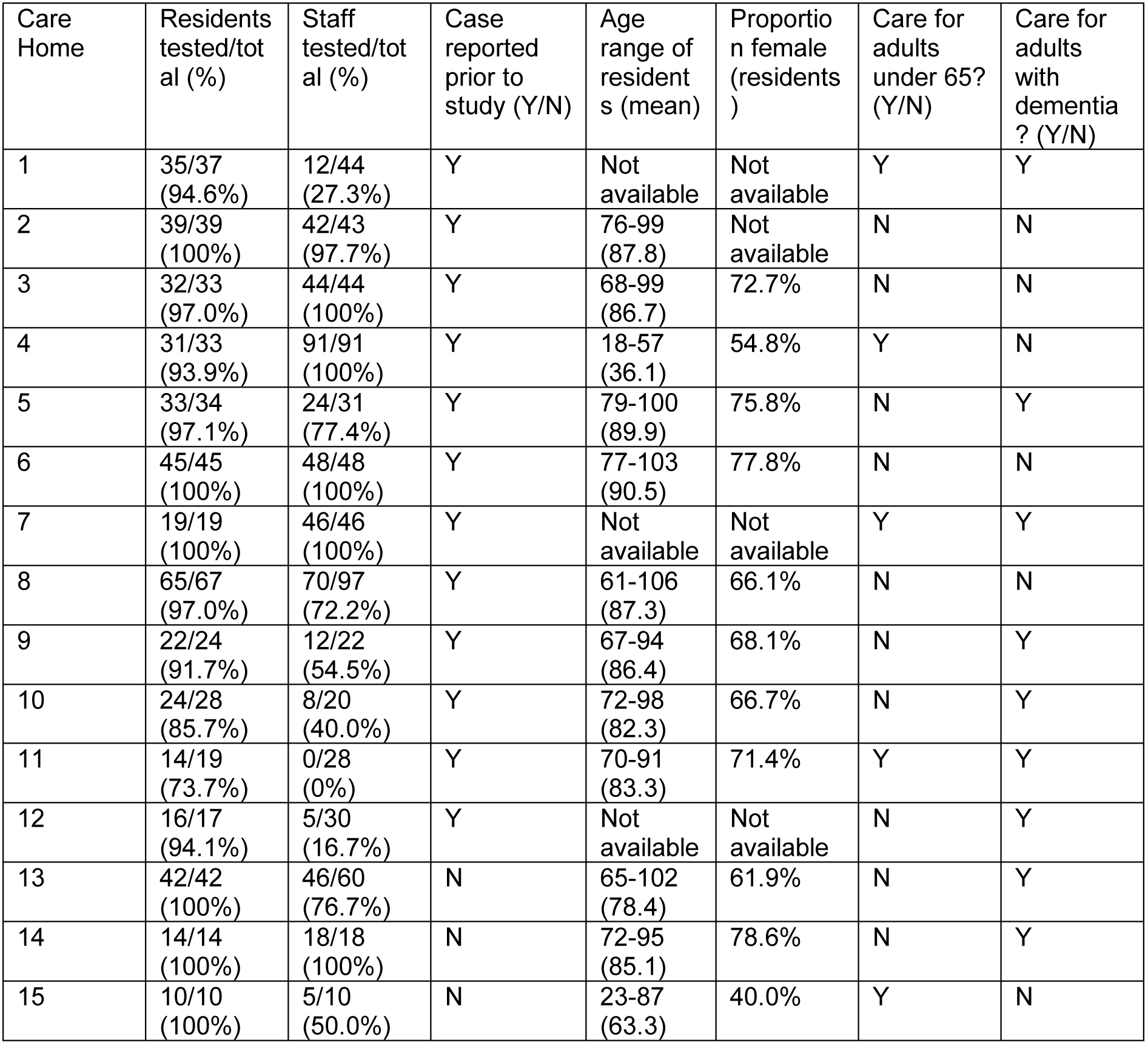
Characteristics of Care homes studied.

Overall, 912 staff and residents were tested for SARS-CoV-2 – 441/461 (95.7%) of residents and 471/632 (74.5%) of staff (Table 1). All care homes except two achieved greater than 90% resident coverage (Table 1). A total of 10 (1.09%) swabs could not be analysed due to labelling or laboratory issues and were reported as void. By contrast, the coverage of staff differed markedly between care homes, from 100% in some homes, to very limited in others (#11, 12 in Table 1).

Overall SARS-CoV-2 was detected in 31 (3.4%) of all tested. Of the 441 tests performed on residents, SARS-CoV-2 was detected in 23 (5.2%). For residents with positive results, 21 (91.3%) were reported by care home managers as being asymptomatic and two (8.7%) as symptomatic at the time of testing (Table 2). Of the 21 residents reported as positive, none went on to develop symptoms in the next two weeks.

**Table 2.**
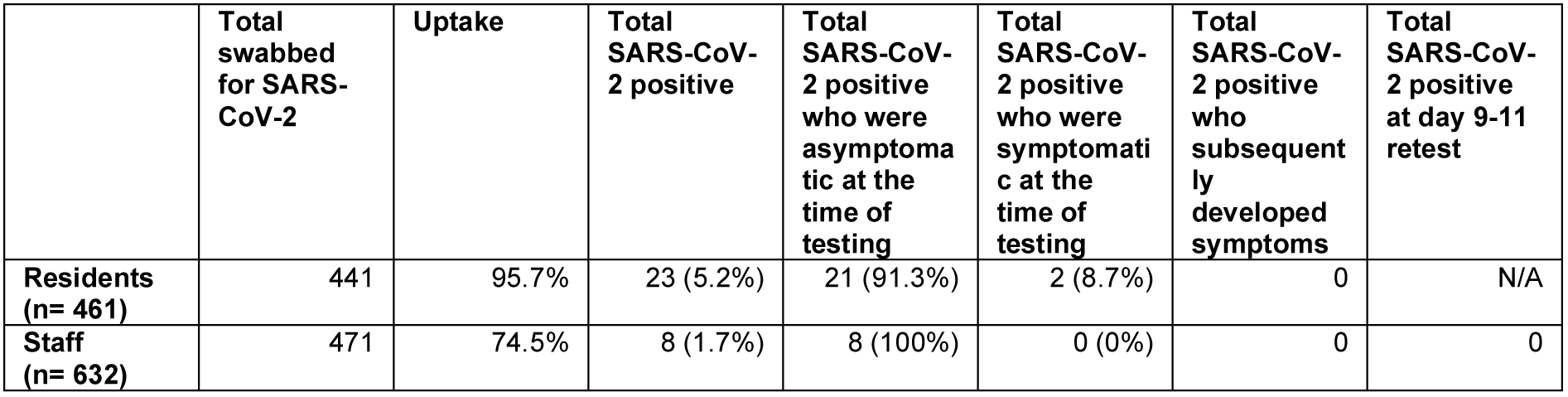
Results of Testing.

Of the 471 tests performed on staff, SARS-CoV-2 was detected in 8 (1.70%). This differs from that in residents (p=0.003). All staff with positive results were reported to be asymptomatic on the day of testing (Table 2), and all were negative on retesting between day nine and eleven.

The three homes that had no suspected or confirmed clinical cases in residents or staff prior to swab-testing taking place had no positive results in staff or residents with staff uptake between 50 and 100%.

## DISCUSSION

In the 15 care homes studied, near complete coverage of residents was achieved (95.7%), with a lower portion of care home staff screened (74.5%, but with wide variation between homes). SARS-CoV-2 was detected in 5.2% of residents, nine out of ten of who were asymptomatic at the time of swabbing, as reported by the care home managers. The point-prevalence of SARS-CoV-2 was lower in care home staff than residents at 1.7% but this may have been affected by the screening policy, which excluded ill staff from attending.

The study found a lower proportion of positive SARS-CoV-2 residents in care homes than in prior descriptive studies in the UK^6^ and United States^11^, but with a higher proportion of those who were detected being reported by care home managers as asymptomatic. The study found a slightly higher prevalence of SARS-COV-2 in residents than in the Whole Care Testing programme with a higher proportion of positive residents being asymptomatic^7^ This may result from different susceptibilities to severe clinical disease in different care homes, differences in infection to testing interval, survivorship bias (that some individuals had died prior to the cross-sectional study) or differences in ascertainment of illness in different homes. As well as the studies taking place at different points in the epidemic.

Care home settings appear to be particularly susceptible to COVID-19 outbreaks. Many residents are vulnerable to severe infection due to their advanced age and multiple co-morbidities and much is still unknown about the transmission of COVID-19 within care homes, despite their vulnerability^11^. In this study, the difference between the proportion of individuals virologically positive (5.2%), and those considered symptomatic by care home managers (~ 0.5%) is striking. This is important because, in the absence of molecular surveillance of SARS-CoV-2 ingress into care homes, public health surveillance has been reliant on symptom-based surveillance and reporting by care home and their managers. This report suggests this activity, formerly a backbone of public health surveillance, may be very insensitive.

As of July 2020, the Department for Health and Social Care announced there will be a roll out of weekly staff testing and testing of residents every 28 day out days in care homes without reported outbreaks. The policy will be reviewed in September 2020. As lockdown restrictions continue to be lifted this study’s findings adds to the evidence for this routine molecular surveillance to be instituted.

## Data Availability

Non-identifiable patient data is available on request.

## Ethics

This study has been subject to an internal ethical and governance review at Public Health England which considered the study design, content and feasibility. The review also covered all legal, financial, regulatory and ethical considerations. As a result of this review, the study was categorized as a public health practice study that was being undertaken to help to manage the COVID 19 outbreak. As no ethical issues were identified it was decided that consideration by an ethics committee would not be necessary

## Patient and Public Involvement

Patients and the public were not involved in the design of this study as it formed part of the local response to the COVID-19 pandemic.

## Transparency Declaration

The lead author affirms that the manuscript is an honest, accurate, and transparent account of the study; no important aspects of the study have been omitted; and any discrepancies from the study as originally planned have been explained.

